# Patterns of GLP-1 Receptor Agonist Initiation Compared With Other Glucose-Lowering and Anti-Obesity Therapies Among Older Adults

**DOI:** 10.64898/2026.09.29.26364340

**Authors:** Julianne D. Brooks, Maria A. Donahue, Aya ElHassan, Jiazi Tian, Sonia Hernandez-Diaz, Deborah Blacker, Hossein Estiri, Jennifer S. Haas, Lidia M.V.R. Moura

## Abstract

**Background:** Glucagon-like peptide-1 receptor agonists (GLP-1RAs) have rapidly expanded from primarily glucose-lowering agents for type 2 diabetes (T2D) to therapies used across diabetes, obesity, and cardiometabolic disease. Prescribing has risen sharply since 2018, especially with the broader uptake of semaglutide-based products. Still, real-world initiation patterns remain incompletely characterized in older adults with varying levels of cognitive vulnerability and frailty.

**Methods:** We analyzed treatment patterns of GLP-1RA use in a retrospective cohort of adults aged ≥65 years with T2D in a regional healthcare system from 2018 to 2026. We describe the demographic and clinical characteristics of new initiators of GLP-1RAs compared with initiators of alternative therapies, including sodium-glucose cotransporter-2 inhibitors (SGLT2i) and dipeptidyl peptidase-4 inhibitors (DPP-4i). We analyzed a sub-cohort of patients to identify the prevalence of baseline cognitive concerns using a large language model (LLM). We used a LASSO logistic regression to identify important predictors of GLP-1RA initiation compared to other therapies.

**Results:** Of 58,754 adults aged 65 years or older with T2D, 5,095 initiated GLP-1RA, 5,483 initiated a SGLT2 inhibitor, and 1,516 initiated a DPP-4 inhibitor. GLP-1RA initiators were younger (mean 75.2 [SD 4.9] vs 77.7 [6.0] and 79.5 [7.1] years) and less often aged 80 years or older (17.7% vs 35.0% and 47.6%). The largest difference across groups was adiposity: mean BMI was 33.8 (6.2) vs 29.7 (5.7) and 28.6 (5.6) kg/m2 (SMD 0.592), with a BMI of 30 kg/m2 or higher in 72.1% vs 41.9% and 34.0%. Glycemic control was similar (mean HbA1c 7.46% vs 7.38% and 7.70%; SMD 0.165). GLP-1RA initiators were younger; more likely to be overweight; had lower average comorbidity index; and less dementia, heart failure, and renal disease than those initiating alternative therapies. GLP-1RA initiators had fewer hospitalizations 1-year prior to drug initiation. In a LASSO model, BMI greater than 27 kg/m^2^ was the strongest predictor of GLP-1RA initiation (positive), followed by baseline congestive heart failure (negative) and baseline hospitalization (negative).

**Conclusion:** In this cohort of older adults, those with T2D who initiated GLP-1RAs were younger and had less comorbidities at baseline compared to initiators of other T2D therapies. Overweight BMI was the strongest predictor of GLP-1RAs initiation, whereas baseline HbA1c was similar across groups, suggesting that the decision to start GLP1-RA is more influenced by adiposity than glycemic control. Our analysis of real-world prescribing patterns appear to be in alignment with guidelines for Type II diabetes treatment^1^, which recommend GLP-1RA treatment for glucose lowering and weight management and SGLT2 treatment in those with risk of cardiovascular disease or heart failure.

## INTRODUCTION

GLP-1 receptor agonists (GLP-1RAs) are central to the treatment of type 2 diabetes (T2D) and are increasingly important for obesity and cardiovascular risk reduction.^2^ Their expansion has been driven by evidence for glycemic improvement, weight loss, and cardiometabolic benefit, alongside a marked increase in prescribing in both diabetic and non-diabetic populations.^3,4^ In patients with T2D and established atherosclerotic cardiovascular disease, including prior stroke, guidelines support the use of a GLP–1RA with demonstrated cardiovascular benefit;^5^ relative reductions in total or nonfatal stroke with GLP–1RA therapy.^6–8^

Real–world GLP–1RA use varies by age, obesity, and diabetes status; comorbid CKD and cardiovascular disease; access-related factors; clinical eligibility, insurance coverage, specialist involvement, and gastrointestinal tolerability further influence treatment selection and persistence.^9–11^ In older adults, additional factors such as frailty, cognitive concerns, caregiver support, and treatment burden may influence whether a GLP-1RA is chosen over an SGLT2 inhibitor, a DPP-4 inhibitor, or another therapy.^12^

Often, real-world prescribing does not align with practice guidelines as there may be delays to adoption and practice change. It is important to understand current prescribing patterns for future studies on real-world comparative effectiveness and safety.^13,14^ Clinicians may preferentially prescribe GLP–1RAs to patients with distinct clinical phenotypes, baseline prognosis, and treatment eligibility; thus, unaddressed treatment selection can produce confounding by indication and channeling bias in observational analyses.^13–15^ Approved GLP-1RA use includes T2D, obesity, and cardiorenal risk reduction; initiation patterns may reflect both clinical risk-based treatment selection and nonclinical factors, including cost, insurance coverage, and prior authorization requirements, and specialty-specific prescribing practices.^16–18^

Evidence on GLP–1RAs in adults aged 65 years and older is expanding, including age-stratified analyses of cardiovascular and kidney outcome trials, as well as emerging real–world observational studies.^19,20^ However, much of the available evidence derives from trial-eligible populations or broad administrative cohorts that incompletely capture frailty, multimorbidity, functional status, and cognitive vulnerability.^21,22^ Real–world uptake of GLP–1RAs and SGLT2 inhibitors remains uneven by age, comorbidity, specialty care, insurance, and sociodemographic characteristics, including among patients with type 2 diabetes and established cardiovascular or kidney risk for whom these therapies may be indicated.^21–23^

This analysis is conducted within an integrated academic medical system in which prescribing patterns may be influenced by centralized formulary decisions, prior-authorization workflows, and specialist-heavy referral pathways. We analyze patterns of GLP-1RA initiation compared with other treatments in a cohort of older adults with T2D, with particular attention to heterogeneity by age, cognitive concerns, frailty, cardiometabolic, cardiovascular, and other comorbidities.

## METHODS

### Study Design

We conducted a cross sectional study describing patterns of new GLP-1RA initiation and alternative therapies in people aged 65 and older with a Type II Diabetes diagnosis from 2018 to 2026. We included patients registered at Massachusetts General Hospital (MGH) or Brigham and Women’s Hospital (BWH) with an encounter with an ICD-10 diagnosis code for Type II Diabetes (E11) between January 1, 2018, and April 1, 2026. We included patients with at least one ambulatory visit in calendar years 2018 and 2019, to select patients who were active in the system during the washout period and at least 1 year thereafter.

### Data Source

We used data from Mass General Brigham (MGB) electronic health records (EHR), which covers 527 clinical locations across Massachusetts and New Hampshire, to identify sampling criteria, baseline variables, and medication initiation. For all covariates, we used a one-year baseline period before medication initiation. Our covariates included: demographics (age, sex, race, ethnicity, language), clinical characteristics (BMI, blood pressure), laboratory measurements (HbA1C, creatinine, eGFR, albumin, LDL), healthcare utilization (outpatient, inpatient, and emergency room visits), polypharmacy (we calculated the number of outpatient medications each individual received during the baseline period using a unique medication identifier and created an indicator variable of whether the patient had more than 10 outpatient medications at baseline) and comorbidities (Charlson comorbidity score, and individually congestive heart failure, renal disease, cerebrovascular disease, stroke). We identified comorbid conditions and calculated the Charlson Comorbidity Index (CCI) using ICD-10 codes from the EHR and the R package ‘comorbidity,’ which includes definitions for dementia, cardiovascular disease, congestive heart failure and renal disease. We identified stroke in the one-year baseline period using encounters with ICD-10 codes for stroke (I60 to I64). We grouped age into 3 categories: 65 to 70, 70 to 80, and 80 years and over. Overweight was defined as a BMI ≥27 kg/m2, and obesity was defined as a BMI ≥30 kg/m2. For vitals and laboratory values, we used the mean if the individual had multiple measurements during the baseline period.

A one-year washout period was applied to exclude individuals with any recorded dispensing or ordering of GLP-1RAs, SGLT2 inhibitors, or DPP-4 inhibitors, ensuring a strict new-user design and reducing prevalent-user bias.

We used medication orders in the EHR to define new drug initiators. We included GLP-1RA and two alternative diabetes therapies: SGLT2 inhibitors and DPP-4 inhibitors (Supplemental Table S1). The medication names included for each class are listed in Supplemental Table S2. We used a 1-year washout to define new initiators.

We analyzed a random sample of 5,000 patients from our cohort to identify cognitive concerns using a previously developed LLM and clinical notes.^24^ We analyzed clinical notes within the one-year baseline period before drug initiation to identify baseline cognitive concerns.

### Statistical Analysis

We used descriptive statistics to characterize the sample. For continuous variables, we used the mean and standard deviation. For categorical variables, we adopted frequencies and proportions. We calculated standardized mean differences (SMD), using a threshold of 0.2 to define a meaningful difference between groups. Missing data were presented in our summary tables and excluded from the model analysis.

We used logistic regression with LASSO to select features associated with medication initiation. We included 35 features in the model, including demographics (age), clinical characteristics (BMI >=27, albumin, A1C, BP, LDL, creatinine, eGFR), baseline healthcare utilization (number of and indicator (Yes/No) of hospitalizations, number of and indicator (Yes/No) of outpatient and emergency department visits within one year before drug initiation), baseline polypharmacy (medication count), baseline comorbidities, and the CCI. We standardized continuous variables to have a mean of zero and standard deviation of one. We used the R package ‘glmnet’ for feature selection, fitting a binomial LASSO logistic regression with alternative therapy as the reference group (GLP1-RA = 1, Other Therapy = 0). We used 10-fold cross-validation to select the largest lambda within one standard error of the minimum. We assessed model performance using the area under the curve (AUC). The analysis was conducted using R Version 2026.01.0+392.

### Data Access and Availability

We take full responsibility for the data, analyses, interpretation, and conduct of the research. We have full access to all the data used for this study. Anonymized data not published within this article will be made available by request from any qualified investigator.

### Standard Protocol Approvals, Registrations, and Patient Consents

This study followed the Strengthening the Reporting of Observational Studies in Epidemiology (STROBE; Table S3)^25^ reporting guidelines and was approved by the Mass General Brigham Institutional Review Board. Because we conducted a secondary analysis of data collected retrospectively, the requirement for informed consent was waived.

## RESULTS

Between 2018 and 2026, 58,754 adults aged 65 years or older had an encounter at MGB with an ICD-10 diagnosis code for T2D (Figure 1). Of these, 40,355 had at least one visit in both the 2018 and 2019 calendar years, ensuring data availability for the washout period and for ascertainment of medication initiation. A further 4,437 were excluded because of a record of a GLP-1RA, SGLT2i, or DPP-4i during the 2018 washout year, leaving 35,918 potential new users. After excluding individuals who contributed to more than one exposure group, the final analytic cohort comprised 12,094 new users: 5,095 GLP-1RA initiators (42.1%), 5,483 SGLT2i initiators (45.3%), and 1,516 DPP-4i initiators (12.5%).

**Figure 1.**
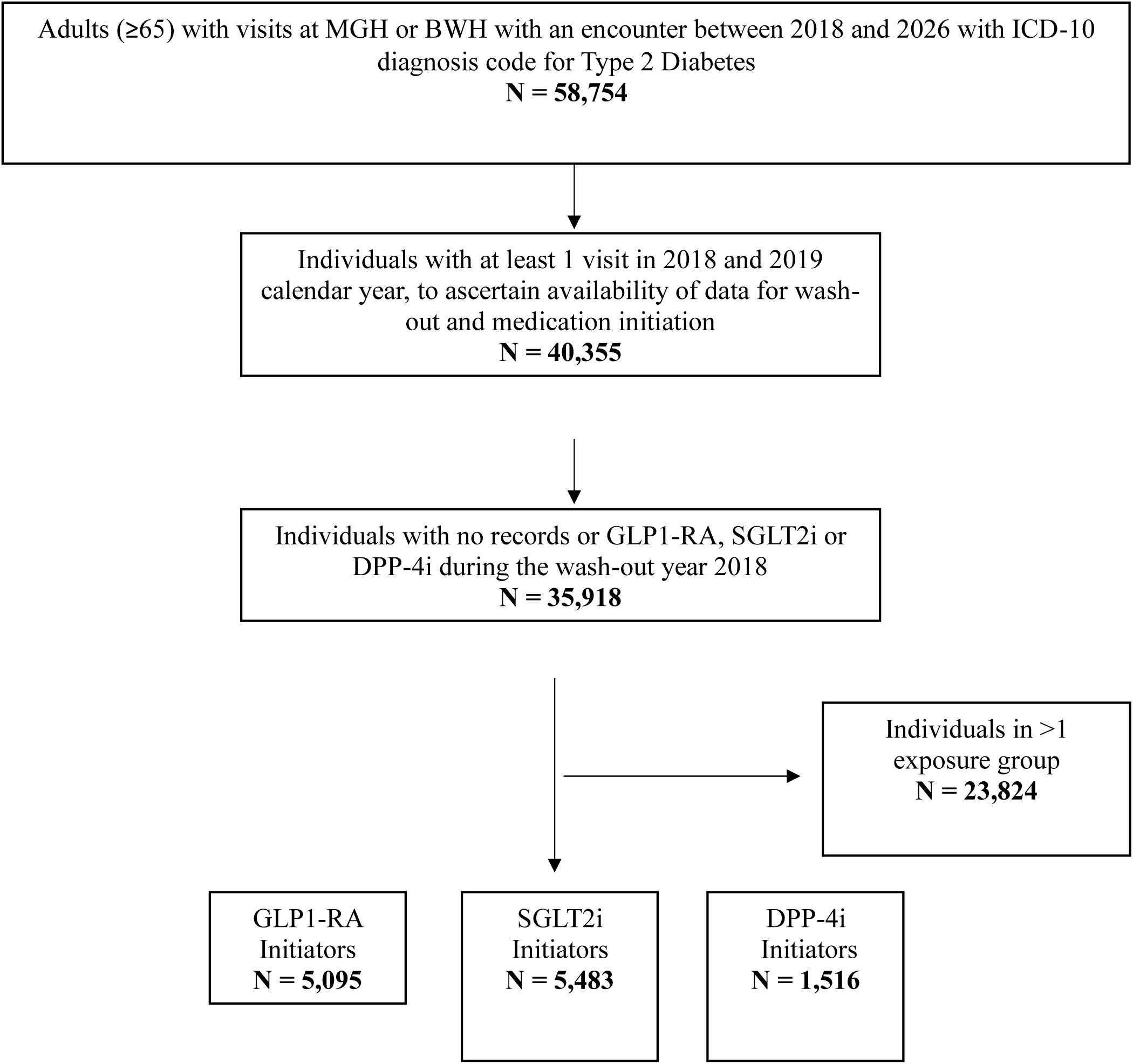

### Demographic Characteristics

Demographic characteristics stratified by exposure group are shown in **Table 1**. GLP-1RA initiators were the youngest of the three groups (mean age 75.2 years, SD 4.9) compared with SGLT2i initiators (77.7, SD 6.0) and DPP-4i initiators (79.5, SD 7.1; p<0.001, SMD 0.478). Age was the second largest difference observed across all baseline characteristics (SMD 0.48). A substantial number of people 80 years and older received GLP-1RA (N = 902), but proportionally there were fewer GLP-1RA initiators in the older age category (17.7% of GLP-1RA initiators were 80 years or older) compared to 35.0% of SGLT2i and 47.6% of DPP-4i initiators. Men comprised 44.4% of GLP-1RA initiators and 43.8% of DPP-4i initiators and 58.9% of SGLT2i initiators (SMD 0.204). Our sample was mostly insured by Medicare (67%) (**Table 1)**.

**Table 1.**
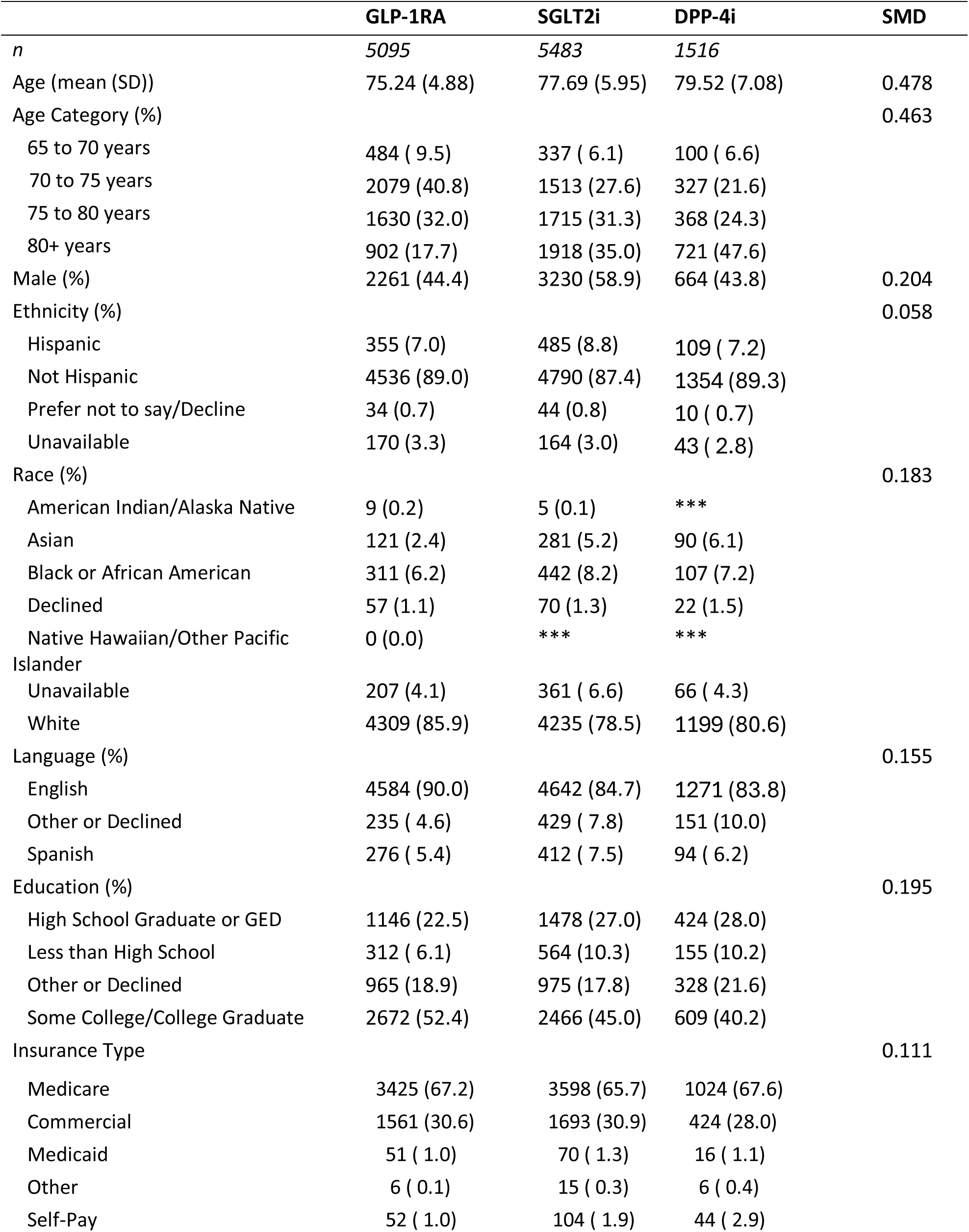
Baseline demographic characteristics of GLP-1RA initiators vs comparator initiators.

### Anthropometric and Metabolic Characteristics

Clinical characteristics of the sample are shown in **Table 2**. BMI was the single characteristic that showed the largest difference between GLP-1RA initiators from comparator initiators (SMD: 0.59). Mean BMI was 33.8 kg/m^2^ (SD 6.2) among GLP-1RA initiators compared to 29.7 kg/m^2^ (SD 5.7) among SGLT2i initiators and 28.6 kg/m2 (SD 5.6) among DPP-4i initiators. A BMI of 27 kg/m^2^ or higher was present in 88.1% of GLP-1RA initiators versus 65.4% and 56.5% of comparator initiators (SMD: 0.50), and a BMI of 30 kg/m^2^ or higher in 72.1% versus 41.9% and 34.0% (SMD: 0.54).

**Table 2.** Laboratory and Vital Signs of GLP-1RA initiators vs comparator initiators.

| Variable (mean (SD)) | GLP-1RA | SGLT2i | DPP-4i | SMD |
| --- | --- | --- | --- | --- |
| *n* | 5095 | 5483 | 1516 |  |
| BMI ≥ 27 | 3879 (88.1) | 3186 (65.4) | 684 (56.5) | 0.497 |
| BMI ≥ 30 | 3176 (72.1) | 2042 (41.9) | 412 (34.0) | 0.542 |
| A1C | 7.46 (1.45) | 7.38 (1.26) | 7.70 (1.30) | 0.165 |
| BMI | 33.81 (6.18) | 29.69 (5.69) | 28.58 (5.63) | 0.592 |
| EGFR | 66.86 (19.92) | 61.43 (19.83) | 57.14 (22.50) | 0.311 |
| Creatinine | 1.08 (0.54) | 1.18 (0.46) | 1.30 (0.85) | 0.226 |
| Albumin | 4.21 (0.37) | 4.16 (0.45) | 4.05 (0.50) | 0.238 |
| Diastolic blood pressure | 72.29 (9.19) | 70.66 (9.65) | 70.56 (9.23) | 0.124 |
| Systolic blood pressure | 132.05 (16.48) | 131.90 (17.75) | 132.03 (16.69) | 0.006 |
| LDL Cholesterol | 69.86 (32.84) | 63.72 (31.49) | 69.76 (33.14) | 0.127 |

Glycemic control, in contrast, was similar across groups. Mean HbA1c was 7.46% (SD 1.45) among GLP-1RA initiators, 7.38% (SD 1.26) among SGLT2 inhibitor initiators, and 7.70% (SD 1.30) among DPP-4 inhibitor initiators (SMD 0.17).

### Renal Function, Lipids, and Blood Pressure

GLP-1RA initiators had better-preserved renal function than either comparator group, with a mean eGFR of 67 (SD 20) for GLP-1RA initiators, 61 (SD 20) for SGLT2i initiators, and 57 (SD 23) for DPP4i initiators (SMD 0.31). The mean creatinine and albumin levels also differed between groups, with SMD values greater than 0.2 (**Table 2**).

### Baseline Comorbidities and Cardiovascular Risk

Baseline comorbidities are shown in **Table 3 and Supplemental Table S3**. A higher percentage of SGLT2i initiators (35%) had congestive heart failure at baseline compared to GLP-1RA initiators (14%) and DPP4i initiators (19%) (SMD: 0.33). Among GLP1-RA initiators, the prevalence of stroke at baseline was 4% compared to 6% for alternative therapies (SMD <0.2). Baseline comorbidity index also varied by treatment group, with values of 3.5 (SD 2.6) for GLP1-RA, 4.4 (SD 2.8) for SGLT2i, and 3.8 (SD 2.9) for DPP4i initiators (SMD: 0.21).

**Table 3.**
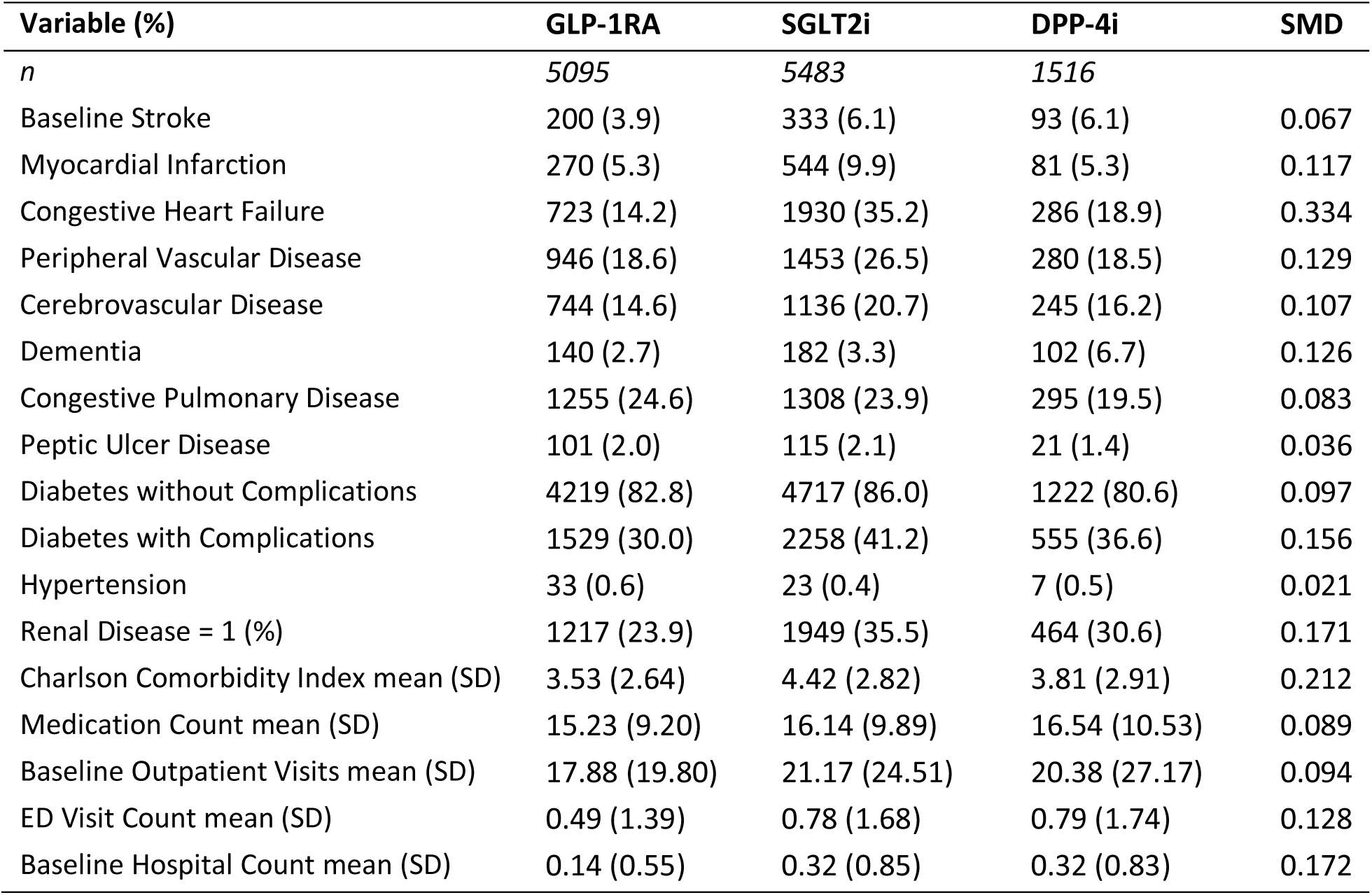
Baseline clinical characteristics of GLP-1RA initiators vs comparator initiators.

| Variable (%) | GLP-1RA | SGLT2i | DPP-4i | SMD |
| --- | --- | --- | --- | --- |
| *n* | 5095 | 5483 | 1516 |  |
| Baseline Stroke | 200 (3.9) | 333 (6.1) | 93 (6.1) | 0.067 |
| Myocardial Infarction | 270 (5.3) | 544 (9.9) | 81 (5.3) | 0.117 |
| Congestive Heart Failure | 723 (14.2) | 1930 (35.2) | 286 (18.9) | 0.334 |
| Peripheral Vascular Disease | 946 (18.6) | 1453 (26.5) | 280 (18.5) | 0.129 |
| Cerebrovascular Disease | 744 (14.6) | 1136 (20.7) | 245 (16.2) | 0.107 |
| Dementia | 140 (2.7) | 182 (3.3) | 102 (6.7) | 0.126 |
| Congestive Pulmonary Disease | 1255 (24.6) | 1308 (23.9) | 295 (19.5) | 0.083 |
| Peptic Ulcer Disease | 101 (2.0) | 115 (2.1) | 21 (1.4) | 0.036 |
| Diabetes without Complications | 4219 (82.8) | 4717 (86.0) | 1222 (80.6) | 0.097 |
| Diabetes with Complications | 1529 (30.0) | 2258 (41.2) | 555 (36.6) | 0.156 |
| Hypertension | 33 (0.6) | 23 (0.4) | 7 (0.5) | 0.021 |
| Renal Disease = 1 (%) | 1217 (23.9) | 1949 (35.5) | 464 (30.6) | 0.171 |
| Charlson Comorbidity Index mean (SD) | 3.53 (2.64) | 4.42 (2.82) | 3.81 (2.91) | 0.212 |
| Medication Count mean (SD) | 15.23 (9.20) | 16.14 (9.89) | 16.54 (10.53) | 0.089 |
| Baseline Outpatient Visits mean (SD) | 17.88 (19.80) | 21.17 (24.51) | 20.38 (27.17) | 0.094 |
| ED Visit Count mean (SD) | 0.49 (1.39) | 0.78 (1.68) | 0.79 (1.74) | 0.128 |
| Baseline Hospital Count mean (SD) | 0.14 (0.55) | 0.32 (0.85) | 0.32 (0.83) | 0.172 |

### Baseline Healthcare Utilization

Baseline healthcare utilization is shown in **Table 3**. The percentage of GLP-1RA initiators with at least one hospitalization during the baseline year was 9.2%. For the other therapies, the baseline hospitalization rate was 19-20%, which is 2 times higher than that of GLP1-RA initiators (SMD: 0.21). A similar pattern was observed for emergency department visits, with baseline percentages of 21% for GLP1-RA, 32% for SGLT2i, and 29% for DPP-4i (SMD: 0.17).

### Baseline Cognitive Concerns

We created a random sample of 5,000 patients to analyze baseline cognitive concerns using an LLM on any available clinical notes. We required that patients had at least one baseline clinical note with more than 100 characters to ensure sufficient text density. We excluded 150 patients who were either A) missing clinical notes or B) only had clinical notes of fewer than 100 characters. In the sub-cohort of 4,850 patients, the overall prevalence of cognitive concerns was 24% (**Table 4**). Among GLP1-RA initiators, 20% had baseline cognitive concerns, and for SGLT2i and DPP-4i, the baseline prevalence was 27% and 32%, respectively.

**Table 4.**
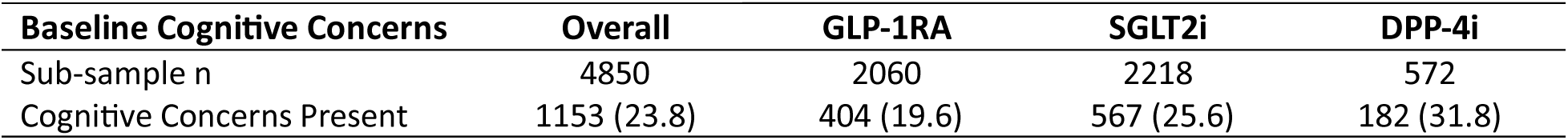
Baseline Cognitive Concerns.

| Baseline Cognitive Concerns | Overall | GLP-1RA | SGLT2i | DPP-4i |
| --- | --- | --- | --- | --- |
| Sub-sample n | 4850 | 2060 | 2218 | 572 |
| Cognitive Concerns Present | 1153 (23.8) | 404 (19.6) | 567 (25.6) | 182 (31.8) |

### Multivariable Predictors of GLP-1RA Initiation

A LASSO model of GLP-1RA initiation versus comparator initiation (**Figure 2**) confirmed that overweight BMI (≥27 kg/m2) was by far the strongest retained predictor. Those with overweight BMI were more likely to initiate GLP-1RA (coefficient of 1.3). Those with congestive heart failure were less likely to initiate GLP-1RA, with a coefficient of −0.80. Hospitalization during the baseline period, age, and diabetes with and without complications were also important negative predictors of GLP1-RA use. Additional predictors are shown in Figure 2. The best performing LASSO regression had an AUC of 0.74 (95% CI: 0.72, 0.76).

**Figure 2.**
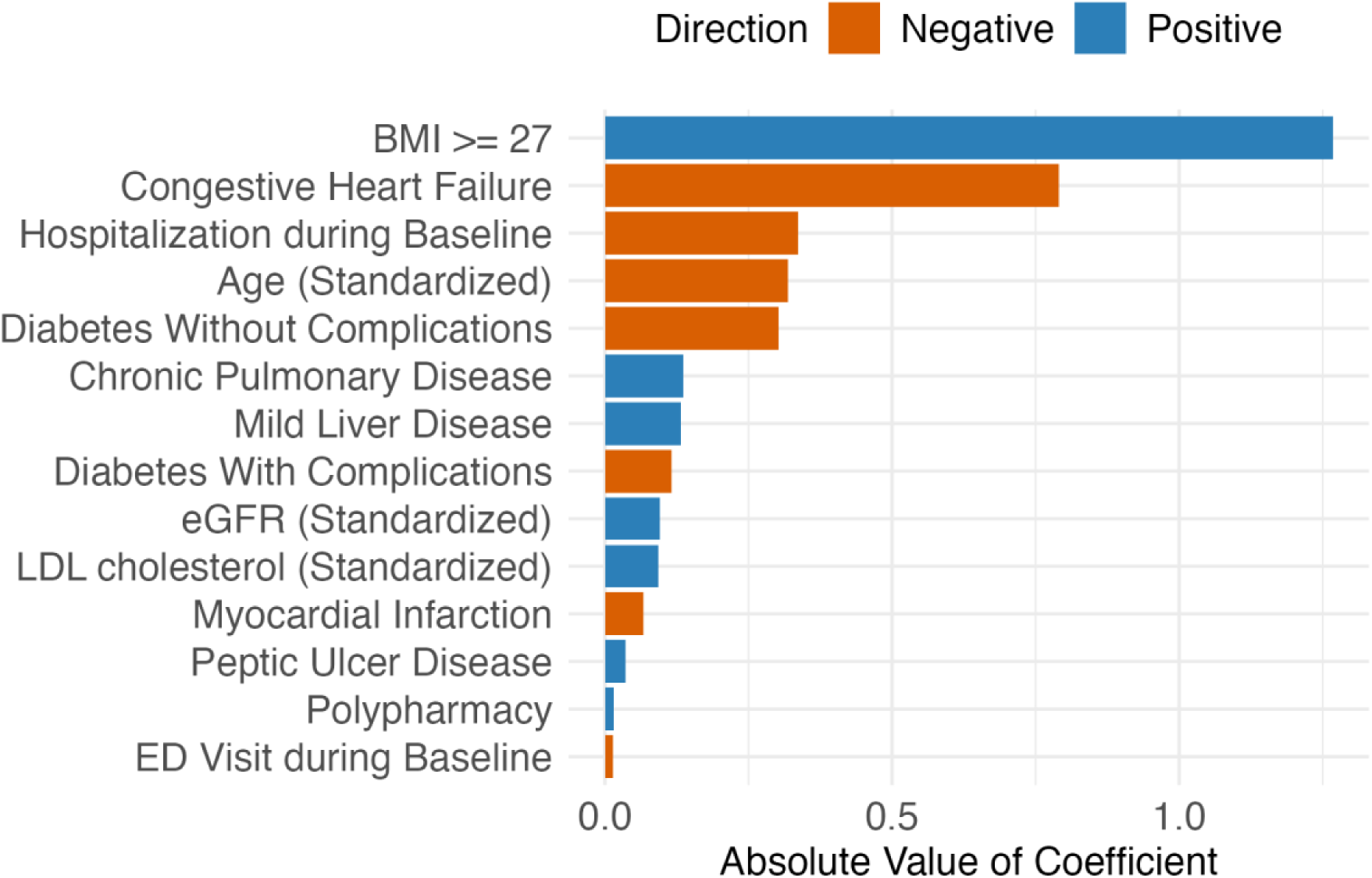
Variable Importance for Predicting GLP1 Use **Legend:** Variable Importance for predicting GLP1-RA use compared to other diabetes treatments. Coefficients were calculated from an LASSO Logistic Regression with 35 features. BMI, Body Mass Index kg/m^2^, eGFR, estimated Glomerular Filtration Rate; LDL, low-density lipoprotein cholesterol; Polypharmacy, 10 or more prescription medications within baseline year

## DISCUSSION

This study characterizes the adoption of GLP-1RAs in routine practice compared with other therapies in a cohort of older adults. Among older adults with T2D at two academic medical centers, GLP-1RAs were preferentially initiated in patients with obesity and comparatively preserved cardiac, renal, and functional status, whereas SGLT2i were initiated in patients with heart failure and chronic kidney disease, and DPP-4i in the oldest patients, who carried the greatest cognitive and comorbidity burden. The model identified obesity as the dominant positive determinant of GLP-1RA initiation and cardiac and acute-care burden as the dominant negative determinants.

GLP–1RA initiation is concentrated among patients with obesity or dual obesity-diabetes indications; this pattern is consistent with the rapid expansion of semaglutide-based prescribing and the marked increase in GLP–1RA use among adults with overweight or obesity without diabetes.^3,26^ On the other hand, patients with chronic kidney disease or cardiovascular disease may indicate a cardiorenal risk-based pattern of adoption, consistent with contemporary diabetes guidelines and evidence from cardiovascular-kidney outcome trials.^27,28^

Lower initiation among the oldest adults or those with documented cognitive concerns could reflect a combination of clinical caution, limited direct trial evidence in very old or frail populations, anticipated gastrointestinal and weight-loss-related tolerability concerns, and access-related administrative burden.^29–32^ If GLP-1RA use increases in these groups, it may indicate broader use among populations underrepresented in prior studies, underscoring the need to monitor safety, treatment persistence, functional outcomes, and quality of life.^29,31,32^

Planned stratification by cognitive concerns and age is important because chronological age alone may not capture the clinical factors to appropriately inform prescribing.^33^ Diabetes guidelines recommend that treatment complexity be matched to an older adult’s cognitive and functional capacity and available caregiver support; documented memory concerns, dementia, caregiver dependence, and functional vulnerability may therefore influence the feasibility and tolerability of injectable therapies requiring dose titration and sustained self-management.^33,34^ This consideration is particularly relevant for GLP–1RAs, for which gastrointestinal adverse effects, dehydration and treatment-related sarcopenia and bone loss can be clinically consequential in older adults with frailty or limited physiologic reserve.^31^ Descriptive analyses incorporating these characteristics may identify clinically meaningful treatment-selection phenotypes that would be hidden in age-only summaries.^30^

Stratification by type 2 diabetes, obesity, chronic kidney disease, and cardiovascular disease is important because GLP–1RAs now have overlapping therapeutic roles.^35–37^ Depending on the patient’s phenotype, the class may be initiated primarily for glycemic management, chronic weight management, reduction of cardiovascular events, or slowing of kidney disease progression.^37,38^ Accordingly, overall GLP–1RA initiation estimates may hide clinically distinct treatment-selection pathways. Recent utilization studies demonstrate that GLP–1RA initiation patterns differ substantially between patients with diabetes and those with overweight or obesity, with marked temporal shifts in the relative contribution and demographic composition of these groups.^3,39^

A major contribution of this descriptive work will be to inform later causal analyses. Significant differences between GLP-1RA initiators and comparator initiators indicate the need for careful active-comparator selection, indication restriction, and subgroup-specific target trial emulation. This concern is now concrete rather than hypothetical. GLP-1RAs were preferentially prescribed for patients with obesity, DPP-4i in older patients with higher HbA1c, more documented dementia, and SGLT2i in patients with substantially more congestive heart failure. Pooled comparison across these groups without careful design would therefore yield severe confounding by indication, treatment tolerance, and underlying frailty.

The results also indicate that no single comparator is adequate across all patient groups. SGLT2 inhibitors were the most clinically similar comparator with respect to glycemic control and completeness of baseline measurement. Still, their strong channeling toward congestive heart failure, which was both the largest comorbidity difference in Table 1 and a dominant predictor in Figure 2, makes them a problematic comparator for any cardiac endpoint. DPP-4 inhibitors were closer to GLP-1RA initiators in sex distribution and in the prevalence of myocardial infarction and peripheral vascular disease, but they were considerably older, more often had documented dementia, and had higher comorbidity, making them a problematic comparator for mortality or any age-sensitive endpoint.

Clinically, a clear description of who initiates GLP-1RAs can help distinguish enthusiasm from equitable uptake. Rapid growth in prescribing does not necessarily mean broad access or appropriate use. Existing evidence suggests substantial discontinuation in older adults and persistent barriers related to coverage, cost, and tolerability, meaning that initiation alone may overstate the degree to which patients are truly receiving sustained benefit from these therapies.^40,41^

Because GLP-1RAs were rarely initiated among patients of advanced age, with documented cognitive impairment, or with high multimorbidity, evidence generation for those most vulnerable to both benefit and harm remains incomplete. The immediate priority is therefore not to evaluate persistence and functional outcomes in groups already receiving these agents, but to determine whether the observed conservatism in the oldest and most vulnerable patients is clinically justified or reflects unwarranted therapeutic inertia or patient refusal to start medications due to cost and to build the evidence base to answer that question.

### Limitations

There are several limitations to our analysis. We analyzed medication initiation from physician orders and did not track whether the medication was covered by insurance, filled at the pharmacy, taken by the individual, or continued. Our analysis only includes prescription orders placed by MGB providers and does not include prescriptions obtained from online retailers. Our sample consists of adults over 65, for whom Medicare is the primary insurer. GLP-1RAs are covered for Medicare beneficiaries with a diagnosis of Type II diabetes (usually with prior authorization); however, prior to 2025, if a beneficiary with traditional medicare coverage (not Medicare advantage) hit a spending limit, they would enter into a coverage gap called the “donut hole” where they would have to pay higher out-of-pocket costs. Starting in 2026, under the GLP-1 Bridge Program, eligible Medicare beneficiaries will pay $50 per month for GLP-1 medications if they are obese or overweight and have other health conditions.

Prescribing patterns observed in this integrated academic network may reflect institutional formulary tiering, prior authorization requirements, and specialist-channeling behavior specific to tertiary care centers. Individuals in our sample may have emergency department visits or hospitalization outside of the MGB system during the baseline period. We included individuals with at least one encounter in 2018 and 2019 to ensure patients had established care within the MGB system. Changes may influence prescribing patterns, availability, and public awareness over time. Community-based or pharmacy-driven prescribing dynamics may exhibit different utilization thresholds.

Exclusion of incomplete records in descriptive and regression analyses assumes missing completely at random; if missingness correlates with higher comorbidity or lower utilization, SMD estimates may be artificially compressed. Additionally, cognitive concern prevalence derived from an LLM on clinical notes is subject to documentation bias (e.g., variable note density by specialty or clinician), and should be interpreted as a proxy for under-documented frailty rather than absolute patient burden.

## CONCLUSION

This descriptive study shows who receives GLP-1RAs among older adults with T2D in routine clinical care. GLP-1RA initiation was driven mainly by adiposity rather than glycemic control. It was less common among patients who were older or had congestive heart failure, kidney disease, documented dementia, or a recent hospitalization. As a result, patients who started GLP-1RAs were generally healthier at baseline than patients in either comparator group. As GLP-1RA use continues to expand and these drugs are increasingly used for different indications, understanding these treatment patterns is important for future causal studies. Comparisons of benefits, harms, and treatment persistence should account for differences in indication, BMI, frailty, and recent acute-care use.

## Disclosures

Authors M.A.D., J.D.B., J.T., D.B., H.E. and J.S.H. have no conflict of interest. L.M.V.R.M. receives support from the Epilepsy Foundation of America and reports no conflict of interest.

## Data Availability

Anonymized data not published within this article will be made available by request from any qualified investigator.

## Notes

### Author Declarations

This study was approved by the Mass General Brigham Institutional Review Board. Because we conducted a secondary analysis of data collected retrospectively, the requirement for informed consent was waived.

### Funding

- NIH: 5R01AG073410-05

